# Assessment of The Knowledge and Abuse of Abortifacient Drugs Among Students in Higher Learning Institutions in Dodoma City

**DOI:** 10.1101/2025.08.12.25332942

**Authors:** Tienyi Mnyoro Daniel, Juma Ally Hamza, Dennis Francis Mrosso

## Abstract

**Aim:** This study assessed the knowledge and abuse of abortifacient drugs among students in higher learning institutions in Dodoma urban district.

**Method:** A cross-sectional study was conducted involving 100 participants who were students from higher learning institutions in the Dodoma urban district employing simple random sampling technique as a participant recruitment method. The data was collected by the use of both interviewers and self-administered questionnaires. Data obtained were analyzed using SPSS version 20, and the results are presented in the figure and tables.

**Results:** Among 100 participants, results showed that 28% (n=28) have used abortifacients, and the majority,while among the 28 respondents who said they have used abortifacients, the majority of them, 89% (n=25), said they have used pills, and 11% (n=3) used herbs. Also, results from this study show that more than half (55%) of the study participants reported that misoprostol, mifepristone (19%), and comb pack (17%) are the commonly used abortifacient drugs in the environment, although pregnancy termination is restricted by laws in Tanzania.

**Discussion:** The use of abortifacients is on increase in higher learning institutions in Tanzania. Most reasons that were given by the respondents for increased abuse of abortifacients were little education on reproductive health, sloppy sex, pressure from partner or friends, fear of stigmatization and an easy accessibility of these abortifacient agents.

## Background

An abortifacient is any substance that is used to terminate a pregnancy. Abortifacients can be pharmaceutical (pills) or herbal. Common pharmaceutical abortifacients are mifepristone and misoprostol. In addition, there are several herbal mixtures with abortifacient claims; however, there is no available data on the efficacy of these plants in humans.

The majority of students who join universities in Tanzania are aged between 19 and 29 years. Most of the female students are enrolled in university at their young age; this exposes them to unplanned and unprotected sexual intercourse, leading to unintended pregnancies, abortions, and sexually transmitted infections. The increased sexual risk behaviors of female university students have been attributed to movement from a restricted rural to a more liberal urban environment, age, and marital status.

Despite the social and cultural importance of childbearing in many African communities, unwanted pregnancies are the source of problems in the families. This is more severe for young girls who often fall pregnant out of wedlock. The best option for them is to go for an abortion just to avoid facing the judgment from their families and the community in general’

Adolescents aged 15-19 years ‘account for 25% of all the unsafe abortions in Africa, according to women in developing countries who were admitted to the hospital for the treatment of unsafe abortion complications. 38-68% were under the age of 20 years. These complications include cervical or vaginal lacerations, sepsis, hemorrhage, bowel or uterine perforation, tetanus, pelvic infections or abscesses, chronic pelvic inflammatory disease, and secondary infertility (Olukoyo et al., 2001). For Tanzania, 23% of all maternal deaths are among young pregnant girls according to UNFPA Tanzania.

The major consequence of unwanted pregnancy worldwide is induced abortion. This can be performed in health care services where by the abortion is provided by a skilled health care provider with proper equipment, including the use of abortifacients and the correct technique, as well as under a sterile environment as part of the reproductive health services. (Ahmed et al., 2021)

According to WHO standards, these services are provided to any woman in need of the services in countries where abortion is legalized, but for the countries where abortion is illegal except under life-threatening conditions (medical grounds), the majority of the women go for unsafe abortions despite all the complications that may arise, ranging from immediate to lifelong complications and even deaths. Adolescents have the highest risk of serious complications from the unsafe abortion; this is worse in countries where abortion is restricted by law. It is estimated that 46 million abortions are performed each year, 20 million of which occur in countries where abortion is prohibited by law (WHO 2006).

Globally, maternal deaths due to complications of unsafe abortion are on the rise from 13 to 20% (WHO 2003; World Health Report 2005). Women are still risking their lives as well as criminal consequences to terminate their pregnancies; this explains their need to avoid or delay having a child at a particular time in their lives. For Tanzania specifically, considering the low contraceptive rate (34%) and the restricted law on abortion, addressing the problem of unwanted and induced abortion, especially among the youths, will help in dealing directly with maternal mortality and morbidity. This was achieved by exploring the extent of unwanted pregnancies and induced abortions among youths, together with their knowledge and practice of contraception, as well as assessing their sexual behavior in general. Therefore, this study aimed at exploring knowledge and abuse of abortifacient drugs among students in higher learning institutions in Dodoma urban district

## Methodology

### Study design and setting

This was an analytical cross-sectional study conducted in the Dodoma region. The city has more than ten (10) colleges and 2 universities. The study was conducted among two universities available in Dodoma city, St. John’s University of Tanzania and the University of Dodoma, and two colleges, which were the College of Business Education and City College of Health and Allied Science. The Dodoma region is located at latitude -60°00’0.00” S and longitude 360°00’0.00” E. The Dodoma region lies in the center of Tanzania in the eastern central part of the country, the main city being about 480 km from the coast, and it covers an area of 41,311 km²with a population of 2,083,588.

The region is administratively divided into seven districts, which are Bahi, Chamwino, Chemba, Dodoma, Kondoa, Kongwa, and Mpwapwa. The region is bordered by the Manyara region to the north, the Tanga region to the northeast, the Singida region to the west, the Iringa region to the south, and the Morogoro region to the east and southeast. The region produces grapes, beans, seeds, grain, and peanuts, and baobao are also raised and marketed.

The study was carried out in Dodoma Urban District (Dodoma Municipal Council), located at 6.1904°S, 35.7407°E, whereby it’s bordered to the west by Bahi District and to the east by Chamwino District, and the population is 410,956. Its administrative seat is the city of Dodoma

### Study population

The targeted population for this study was the students among higher learning institutions available in Dodoma City who consented to be participants in the study; participants who did not consent were excluded from the study.

### Data Collection Tool

Data was collected using a semi-structured questionnaire with open- and closed-ended questions. The questionnaire was translated from English into Swahili and then back into English to check for consistency. Swahili is the native language of Tanzanians; hence, all questionnaires were translated accordingly. The questionnaire consists of socio-demographic characteristics of respondents, history of pregnancies and induced abortion, knowledge about use and effects of abortifacients, and other factors influencing and related to unwanted pregnancies and induced abortion among students of higher learning institutions in Dodoma city. A pilot study was undertaken to assess the accuracy of the data-gathering tools in order to assure the tool’s validity.

### Sample size determination

The sample size for the study was computed by using Cochran’s formula.Therefore, the level of confidence of 95% and margin of error 0.05 was used in this study. Based on this, the maximum sample size was calculated as follows:

From the **Cochran’s formula**

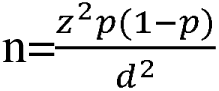

n= Sample size required

z= Standard normal deviate corresponding to significance level of 5% (95% C.I = 1.96)

d= Marginal error set 5%.

p= Prevalence of induced abortion (15%).

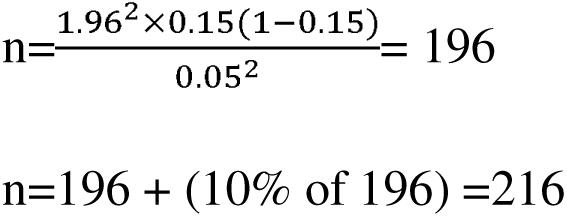

By adding non response rate 10% the sample size becomes 216; because of the limited time and resources 100 respondents was used as a desired sample size.

### Sampling procedure

A sampling technique that was used is a simple random sampling method of the respondents who were available at the right place and at right time who were being informed about the objectives of the study and given and sign informed consent were recruited as participants of the study while those who did not give consent, those who were sick, those who were having university examinations and those who were out of the University campus were excluded from the study.

### Data analysis

The data were coded, checked, and processed with SPSS version 20 so as to generate descriptive statistical information to be presented in form of tables, figures, pie-charts and bar graphs.

### Ethical considerations

Ethical clearance was obtained from the School of Pharmacy at St. John’s University of Tanzania, and all relevant ethical and legal considerations that are applicable to scientific research were also obtained, as well as permission from the authority in higher learning institutions to conduct a study.

Individual consent for the participants was guaranteed, and confidentiality was maintained. The participants have to be informed about how the study was conducted and the purpose of the study. Only those who were willing to participate were enrolled, and every participant was free to respond or not to respond to questions.

No disclosure of information that was obtained from respondents without the permission from the respondents. No pictures, names, or phone numbers of participants were recorded on the questionnaire or elsewhere to ensure confidentiality.

Subjects’ personal privacy was protected, and confidentiality of data was maintained by making sure the respondents remained anonymous. All the information that was gathered from them was only to be used for the purpose indicated previously during permission seeking.

## Results

### Demographic information

From the total of 100 students of higher learning institutions majority (55% n=55) were aged 21-25, (21% n=21). On gender majority 63% (n=63) of the participants were of the participants were female while majority (55%, n=55) of the respondents’ educational level were degree while majority of the respondents 72% n=72) were unmarried as shown in table 1.

**Table 1:**
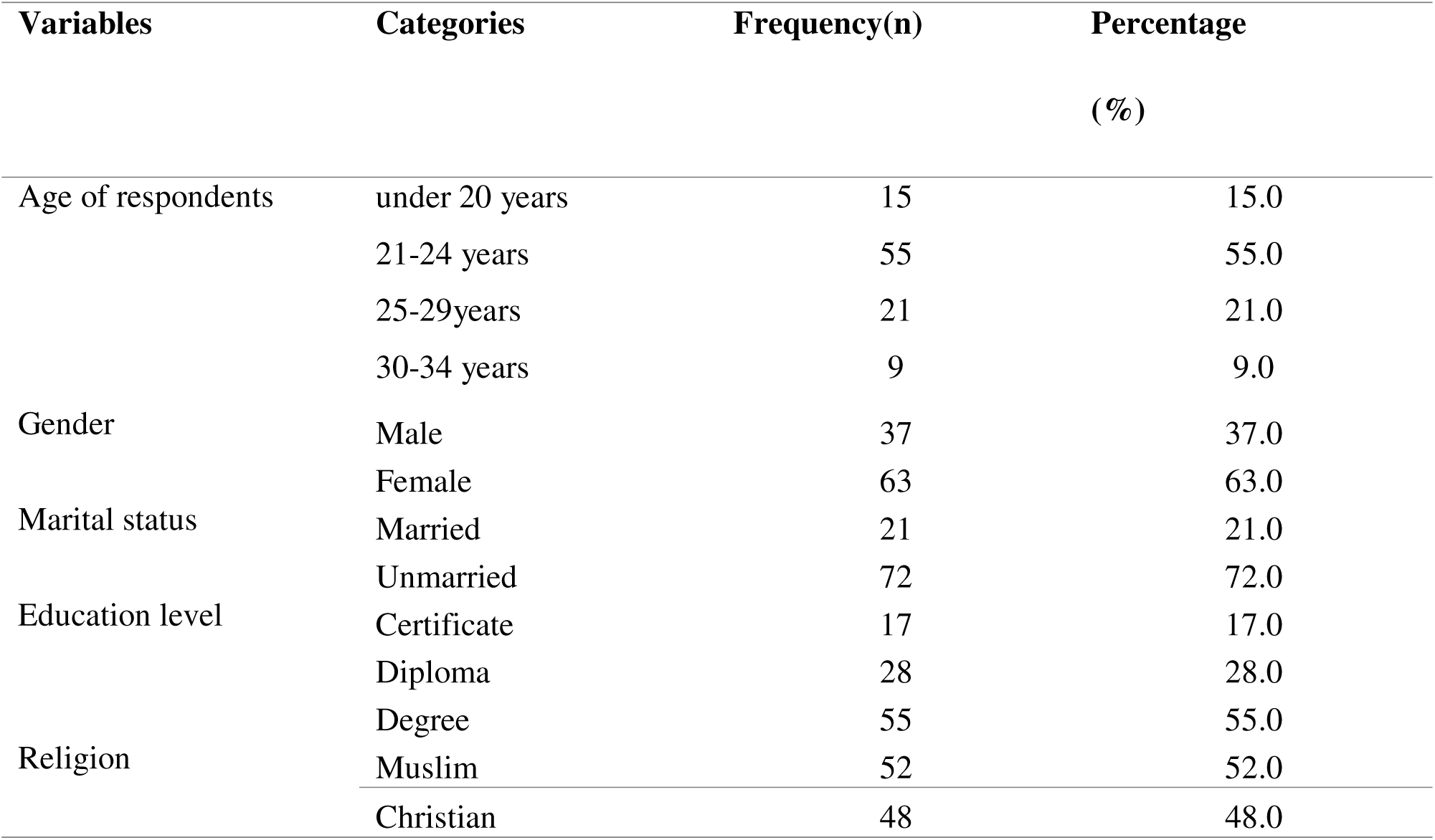
Demographic information.

| Variables | Categories | Frequency(n) | Percentage<br>(%) |
| --- | --- | --- | --- |
| Age of respondents | under 20 years | 15 | 15.0 |
|  | 21-24 years | 55 | 55.0 |
|  | 25-29years | 21 | 21.0 |
|  | 30-34 years | 9 | 9.0 |
| Gender | Male | 37 | 37.0 |
|  | Female | 63 | 63.0 |
| Marital status | Married | 21 | 21.0 |
|  | Unmarried | 72 | 72.0 |
| Education level | Certificate | 17 | 17.0 |
|  | Diploma | 28 | 28.0 |
|  | Degree | 55 | 55.0 |
| Religion | Muslim | 52 | 52.0 |
|  | Christian | 48 | 48.0 |

### Prevalence and types of abortifacients abuse

Among the study participants, only 28% (n=28) have used abortifacients, with 89% (n=25) of them reporting using pills and 11% (n=3) using herbs. Also, results from this study show that more than half (60%) (n=55) of the study participants reported misoprostol as the most commonly used abortifacient drug in the environment, as shown in table 2.

**Table 2:** Commonly used and types of abortifacient agents.

| Variables | Categories | Frequency(n) | Percentage (%) |
| --- | --- | --- | --- |
| Have you ever used abortifacient? | Yes | 28 | 28.0 |
|  | No | 72 | 72.0 |
| How many times have you used abortifacient | 1 | 18 | 18.0 |
|  | 2 | 8 | 8.0 |
|  | 3 | 2 | 2.0 |
| Which types of abortifacients have you used? | Pills | 25 | 89 |
|  | Herbs | 3 | 11 |
| What are the common used abortifacient drugs in your environment | Misoprostol | 55 | 60.0 |
|  | Mifepristone | 19 | 21.0 |
|  | Comb pack | 17 | 19.0 |

**Table 3:**
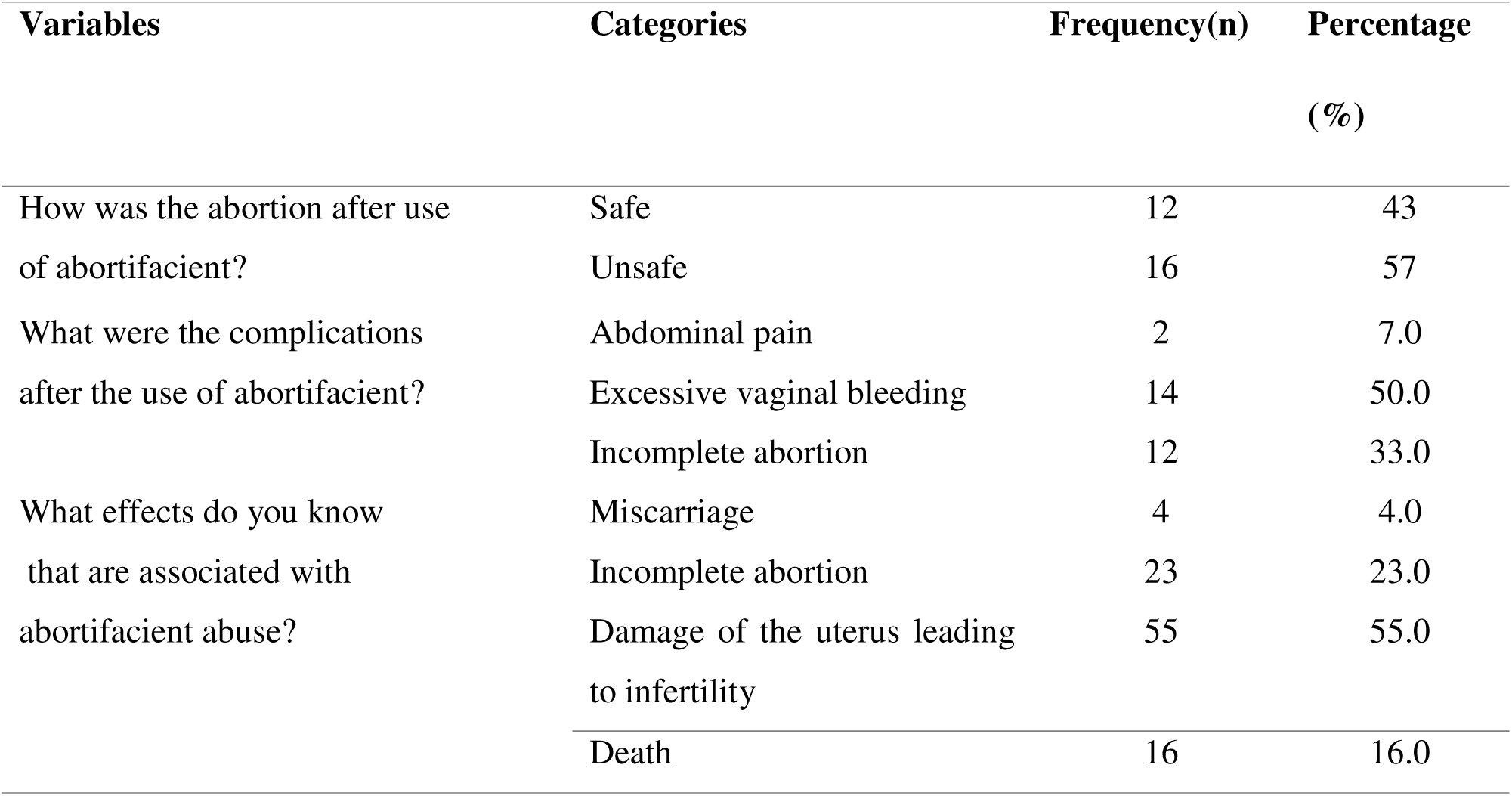
Most commonly short term and long-term effects associated with abortifacient abuse.

| Variables | Categories | Frequency(n) | Percentage (%) |
| --- | --- | --- | --- |
| How was the abortion after use of abortifacient? | Safe | 12 | 43 |
|  | Unsafe | 16 | 57 |
| What were the complications after the use of abortifacient? | Abdominal pain | 2 | 7.0 |
|  | Excessive vaginal bleeding | 14 | 50.0 |
|  | Incomplete abortion | 12 | 33.0 |
| What effects do you know that are associated with abortifacient abuse? | Miscarriage | 4 | 4.0 |
|  | Incomplete abortion | 23 | 23.0 |
|  | Damage of the uterus leading to infertility | 55 | 55.0 |
|  | Death | 16 | 16.0 |

**Table 4:** Factors contributing to abuse of abortifacients.

| Variables | Categories | Frequency(n) | Percentage (%) |
| --- | --- | --- | --- |
| What are the reason(s) for high abuse of abortifacients among students of higher learning institutions | Little education on reproductive health | 10 | 10.0 |
|  | Sloppy sex | 20 | 20.0 |
|  | Fear of stigmatization | 3 | 3.0 |
|  | Pressure from partner or friends | 11 | 11.0 |
|  | Easy access of abortifacients | 1 | 1.0 |
|  | All are the reasons | 55 | 55.0 |

**Figure1:**
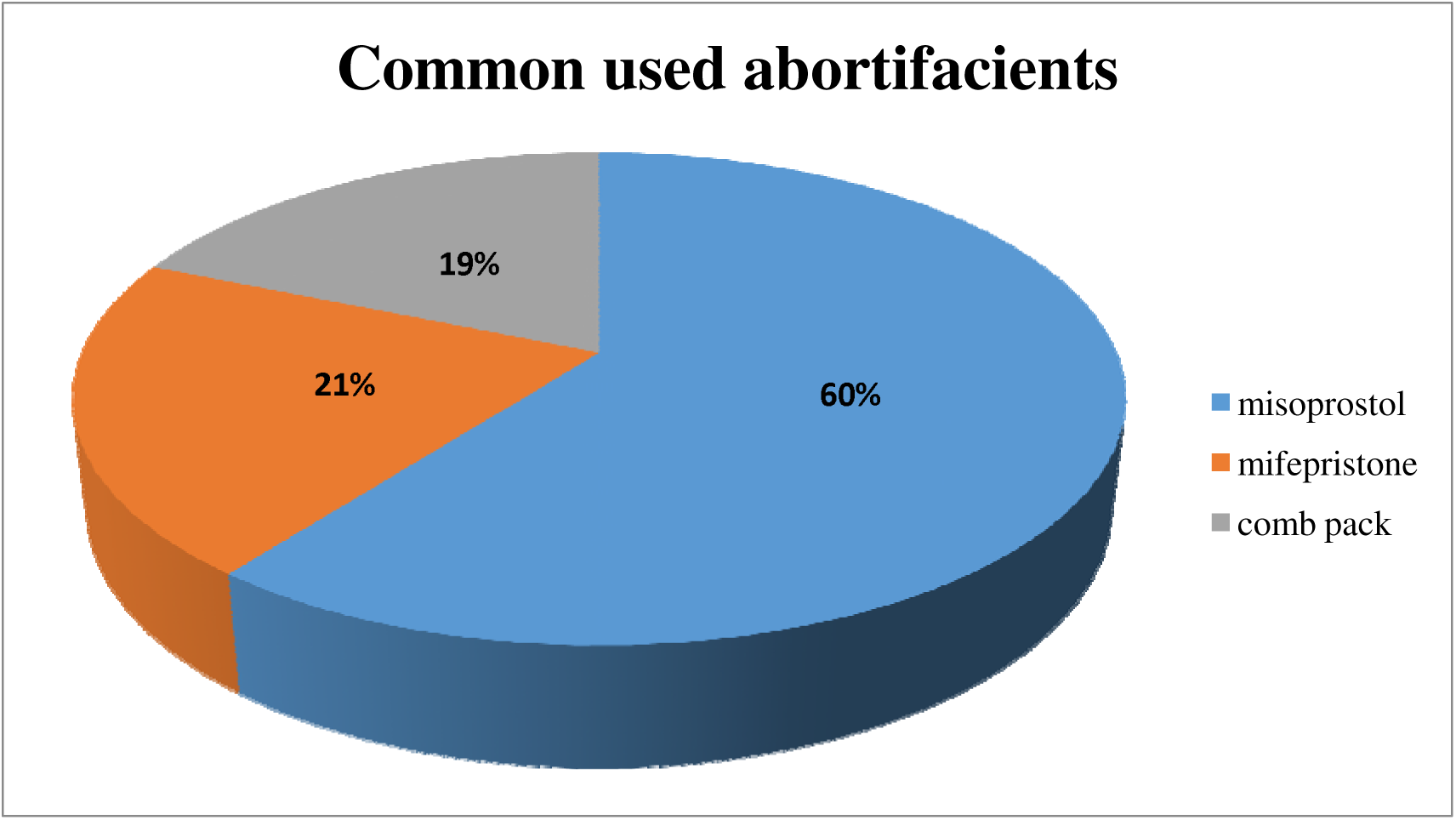
Common abused abortifacients drugs

### Short term and long-term effects associated with abortifacient abuse

Of the 28 (28%) respondents who reported using an abortifacient, 16 (57%) reported it to be unsafe. Twelve (43%) of the respondents reported an incomplete abortion, 2(7%), and 14(50%) reported heavy vaginal bleeding as side effects of using an abortifacient. When asked about the most frequent short- and long-term consequences of abortifacient abuse, the majority of respondents 55(55%) reported that the abuse caused damage to the uterus, which resulted in infertility; 23(23%) said that it caused an incomplete abortion; 16(16%), said that it caused death; and 4(4%), said that it caused miscarriage.

### Factors contributing to abuse of abortifacients

Because 20% of respondents cited sloppy sex as the primary cause of the high rate of abortifacient abuse, the results also highlight potential contributing factors where 11% cited peer or partner pressure; 10% lack knowledge about reproductive health; 3% are afraid of being stigmatized, and 1% have easy access to abortion pills. However, the majority of respondents 55 (55%) stated that all of the factors may lead to abortifacient abuse.

**Figure 2:**
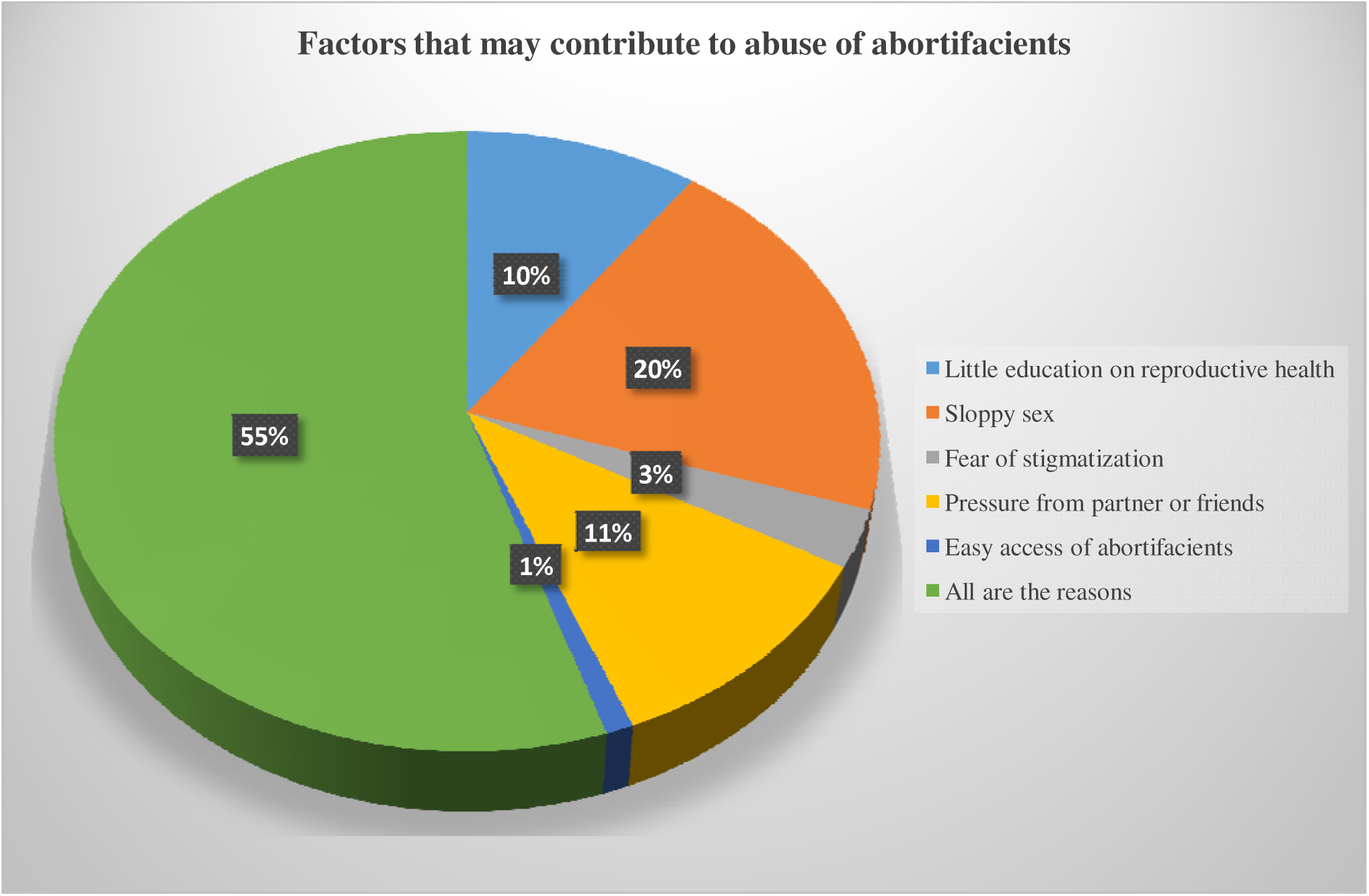
Factors that may contribute to abuse of abortifacients

### Awareness among students on policies, laws and regulations toward abuse of abortifacients

From the study findings majority of the respondents 70% (n=70) reported it to be illegal as illustrated in table 5.

**Table 5:** Student’s awareness on policies, laws and regulations toward abuse of abortifacients.

| Variables | Categories | Frequency(n) | Percent(%) |
| --- | --- | --- | --- |
| What do current nation laws say about abortifacient abuse | Legal | 9 | 9.0 |
|  | Illegal | 70 | 70.0 |
|  | I don't know | 21 | 21.0 |

**Figure 3:**
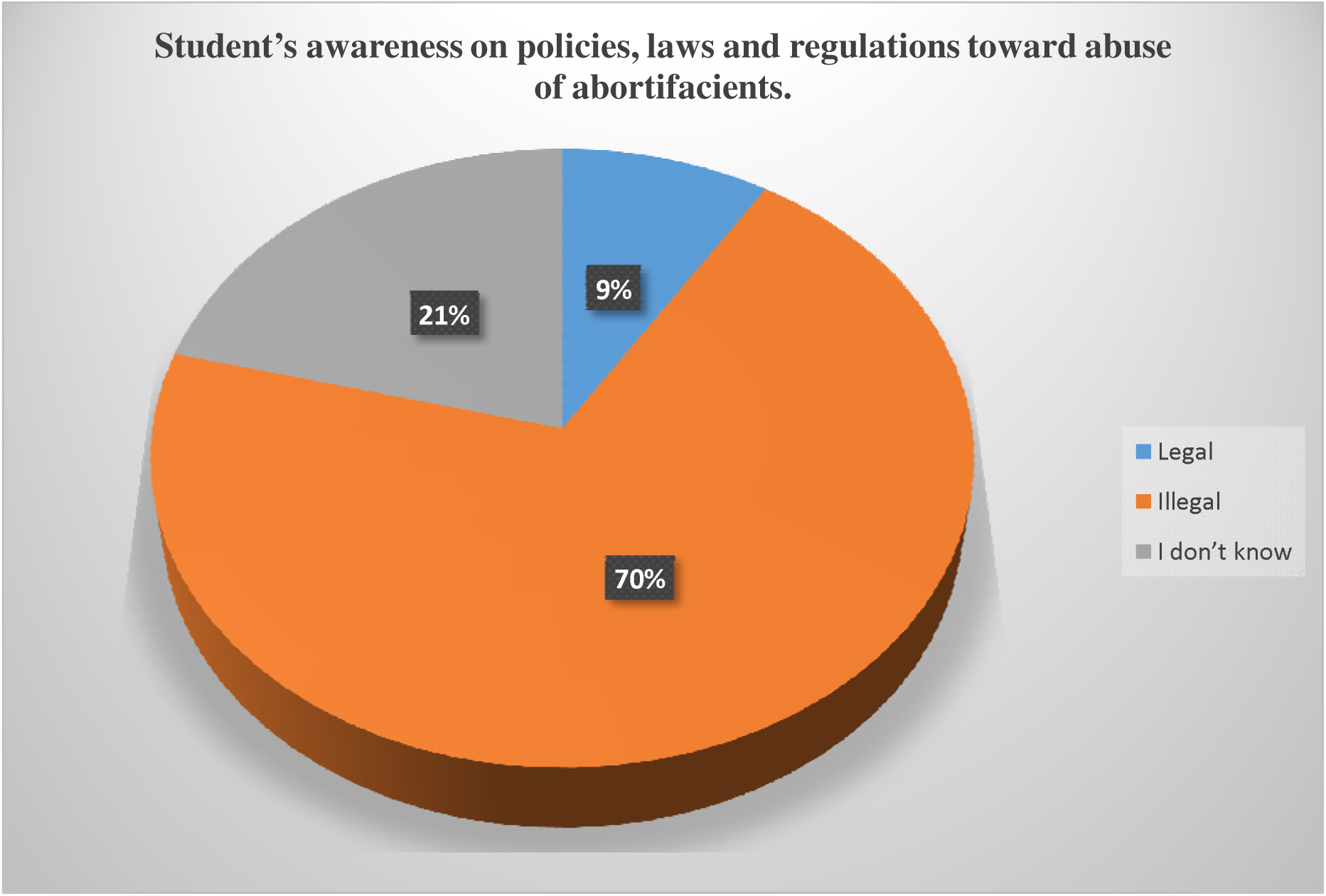
Student’s awareness on policies, laws and regulations toward abuse of abortifacients.

## Discussion

### Prevalence and types of commonly abused abortifacients among students of higher learning institutions

In our study, the use of pills 89% as an abortifacient was reported as the most commonly method used compared to other forms of abortifacient agents such as herbs which was 11%. Other studies elsewhere have revealed similar results where pills were the most used, (Rasch et al., 2019). In addition, only a few indicated that they aborted using herbs. This could be due to the restrictive abortion law in place and negative attitudes towards abortions by health care workers and the fear of being stigmatized, (Cresswell et al., 2016).

Also, results from this study show that more than half 60% of the study participants reported that Misoprostol, Mifepristone 21% and Comb pack 19% as the common used abortifacient drugs around the environment. This is similar to the study done in Ghana on the resolution of unintended pregnancy among female students at university of Ghana on 480 students when asked about their knowledge on the type of drug used Six percent of the students had ever heard about Misoprostol to terminate pregnancy of female students (El-Adas, 2017). And a study done on knowledge and practices among medical abortion seekers in southeastern Nigeria in 100 consecutive abortion seekers from which fifty-five percent of respondents were students where 33% had a tertiary education level 48% had used Misoprostol for pregnancy terminations this is because 73% and 27% of subjects had knowledge of misoprostol and Mifepristone, respectively (Adinma et al. 2019).

### Short term and long term effects associated with abortifacient abuse

Regarding complications, the findings from the study shows majority of respondents 14(50%) reported excessive vaginal bleeding, 12 (43%) incomplete abortion and 2(7%) abdominal pain as were the complications after the use of abortifacient. Also most commonly short term and long term effects associated with abortifacient abuse, majority 55(55%) reported damage of the uterus leading to infertility, 23(23%), 16(16%) said lead to death and 4(4%) responded miscarriage as effects associated with abortifacient abuse.

This is very similar to findings in a study done among university undergraduates in Ibadan, Nigeria, (Cadmus and Owoaje, 2017). The study also revealed that More than half (52.9 %), cited infertility as an adverse outcome of abortifacients. Our study found that 43% had negative attitude(unsafe) towards abortions. In congruence with this, a study in Nigeria reported that attitudes of female students of universities poorly supported unsafe abortions, (Paluku et al., 2020).

### Factors contributing to abuse of abortifacients

There are several reasons for a student to end up having abortifacients for this study it was found that reasons that may contribute to abuse of abortifacients as 20% respondents mentioned sloppy sex as being the major reason of high abuse of abortifacients. 11% said pressure from partner or friends. 10% little education on reproductive health, 3% Fear of stigmatization and 1% Easy access of abortifacients. But the majority of the respondents (55%) said also all the reasons may contribute to abuse of abortifacients.

Our findings are similar to the study conducted in Kenya which reported that abortion rates are higher among university students than other categories, this is because students want to avoid having their educational aspirations terminated. (Stambach 2018). These factors also apply for the rate of induced abortion because by the time student with unwanted pregnancy seeks for medical attention, she has already made her mind and most of the time is to terminate the pregnancy (Baginsk 2017). Studies have found that more than 40% all unsafe abortions in developing countries occur among university student (Baginsk 2017).

Our finding is in contrast with study done in Niger by Baiden, 2019 which reported that many participants admitted that their socio-economic conditions such as financial difficulties, unemployment and inadequate economic support made them indulge in unsafe abortion practices. Many of these women were also in school (students) and were not ready to be mothers. Women who are most vulnerable to unsafe abortions are usually younger, poorer, and lack partner support (Baiden, 2019).

Study conducted in Ethiopia was found that sloppy sex in the youngest age group specifically at college level lead to abortion because students believe they are unable to get pregnant are most likely to conduct induced abortion (Tesfaye et al., 2017).

### Awareness among students on policies, laws and regulations toward abuse of abortifacients

In this study, results show that majority 70% (n=70) of the study participants said it is illegal compared to 9% (n=9) who reported that it is legal and 21(21%) said they don’t know whether is legal or illegal to use abortifacient.

However, our findings are similar to study done by Kavanaugh& Anderson in 2019, that nearly one third of final year students do not agree that abortion should be legally permissible to save the life of the woman. This finding is not only discouraging, but is also inconsistent with the current abortion laws and stands in contrast with attitudinal surveys conducted with health professional in settings where abortion is similarly legally restricted. Further, the overwhelming majority of respondents believe that contraception should not be legally available to unmarried women. Again, this is not consistent with policy or the evidence surrounding contraceptive use. In fact, the evidence demonstrates that contraceptive methods reduce pregnancy-related mortality and morbidity as well as the risk of developing reproductive cancers and can be used to treat many menstrual related symptoms and disorders (Kavanaugh& Anderson 2019). Access to contraception is also a recognized priority in the adolescent reproductive health field and a major pillar of reproductive justice.

A study by Voetagbe et al, 2018 in Ghana found that despite the fact that there is an existing reproductive health policy in Ghana which specifies the need for safe abortion services in Ghanaian health facilities, many students and health providers lack knowledge on the policy. A high proportion of students did not know of the policy governing the provision of safe abortion care in Ghana. It is not surprising that many health providers particularly nurses do not educate women on abortion services in Ghanaian health facilities as part of their reproductive health care for women (Voetagbe et al, 2018).

## Conclusion And Recommendation

### Conclusion

From the study it was found that although pregnancy termination is restricted by law in Tanzania, it is widely practiced and almost always unsafe and contributes to the country’s high maternal morbidity and mortality. Yet the majority of abortion-related deaths are preventable, as are the unintended pregnancies associated with abortion. The use of abortifacients is on increase in higher learning institutions in Dodoma city. Most reasons that were given by the students for having unwanted pregnancies and induced abortion were that they were still in school and they don’t have enough money to take care of the baby respectively. Also it was found that abortifacients abuse results in to different health effects which include incomplete abortion, infections, infertility and it can also lead to death.

### Recommendation

➢ Ministry of Health, Community Development, Gender, Elderly and Children (MoHCDGEC) in partnership with other implementing partners, local and international to focus their strategies on the need to prevent unintended pregnancies and unsafe abortions through the implementation of supportive policies that enhance commitment to the provision of comprehensive sexual education in tertiary institutions in Tanzania.
➢ Giving emphasis on the promotion of birth control in order to avoid abortion and implementation of supportive laws and policies that prevent irrational use and easy access of these abortifacient drugs in the community.
➢ Ministry of education should incorporate awareness of induced abortion and its effects among students in the curriculum of secondary education level this contributes to more awareness regarding induced abortion and its effects to students especially female of all ages to decrease the prevalence of induced abortion in the country.
➢ Further studies should be conducted to assess knowledge, and abuse of abortifacients to other education levels apart from higher learning institutions such as secondary schools.

## Data Availability

All data produced in the present study are available upon reasonable request to the authors

---

**Figure 1:**
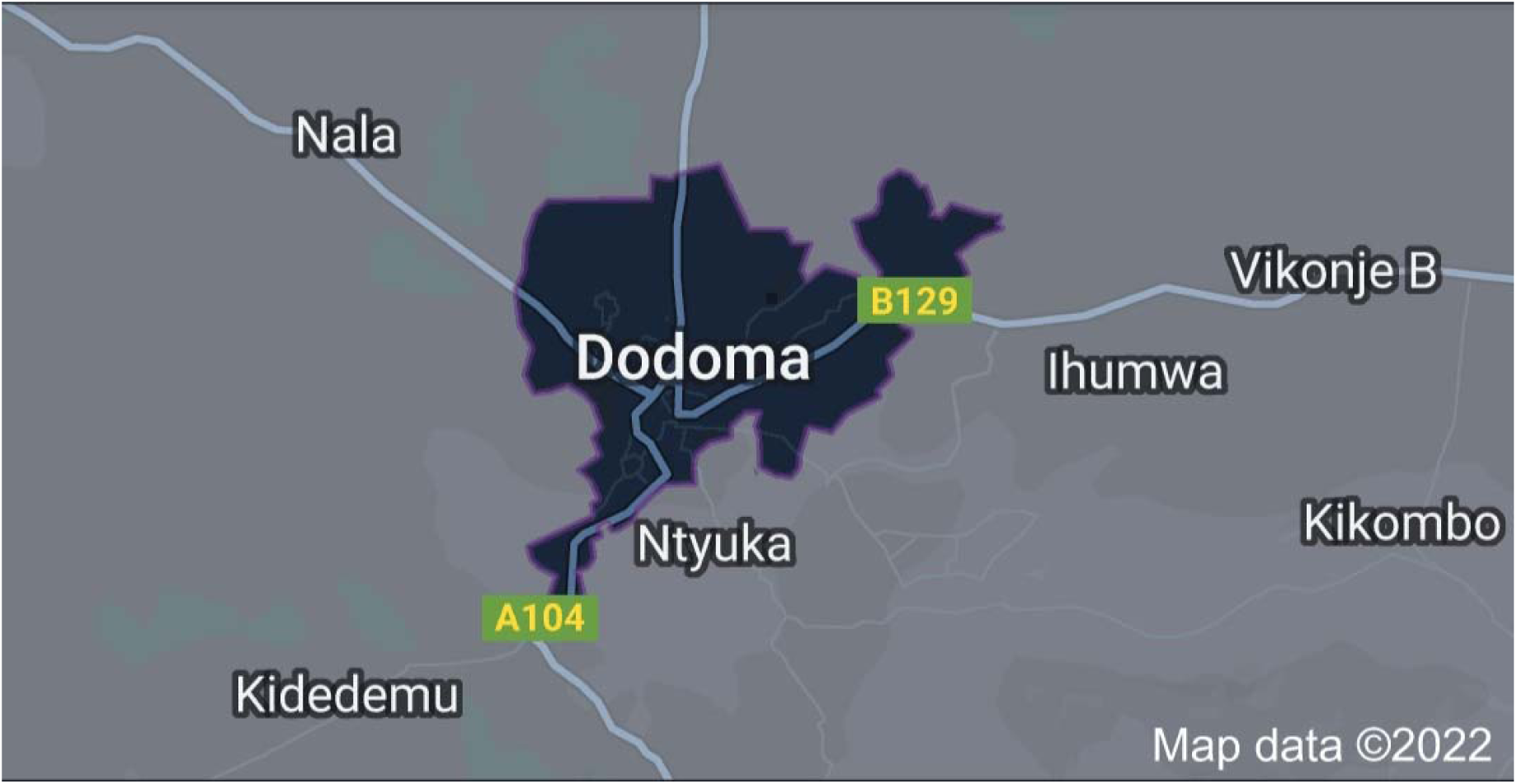
A map of Dodoma city: http://maps.app.goo.gl/EvubW1L1aXKoQW9F9

## RESEARCH ETHICAL CLEARANCE CERTIFICATE

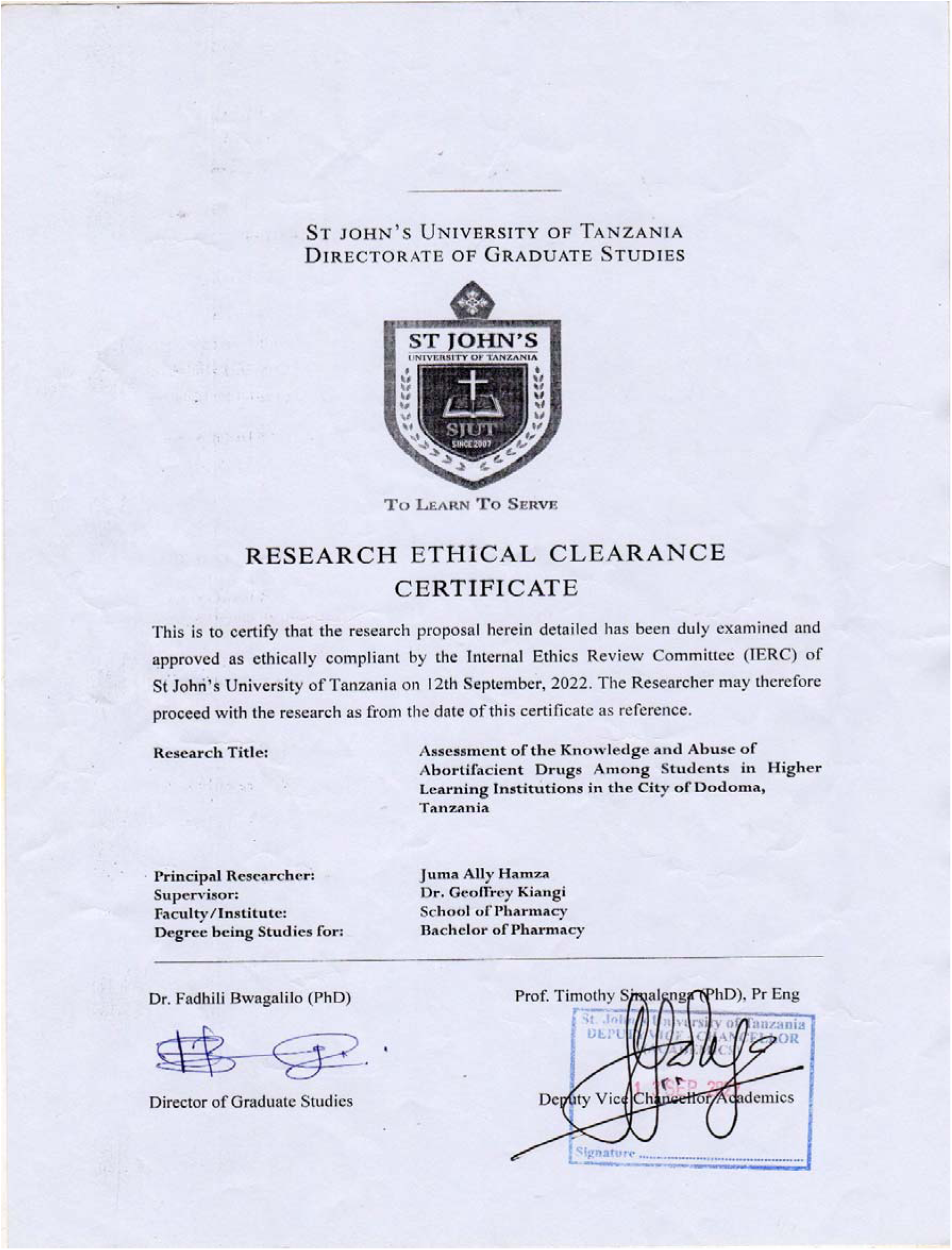

